# Digitally Diagnosing Multiple Developmental Delays using Crowdsourcing fused with Machine Learning: A Research Protocol

**DOI:** 10.1101/2023.03.05.23286817

**Authors:** Peter Washington

## Abstract

**Background:** Roughly 17% percent of minors in the United States aged 3 through 17 years have a diagnosis of one or more developmental or psychiatric conditions, with the true prevalence likely being higher due to underdiagnosis in rural areas and for minority populations. Unfortunately, timely diagnostic services are inaccessible to a large portion of the United States and global population due to cost, distance, and clinician availability. Digital phenotyping tools have the potential to shorten the time-to-diagnosis and to bring diagnostic services to more people by enabling accessible evaluations. While automated machine learning (ML) approaches for detection of pediatric psychiatry conditions have garnered increased research attention in recent years, existing approaches use a limited set of social features for the prediction task and focus on a single binary prediction.

**Objective:** I propose the development of a gamified web system for data collection followed by a fusion of novel crowdsourcing algorithms with machine learning behavioral feature extraction approaches to simultaneously predict diagnoses of Autism Spectrum Disorder (ASD) and Attention-Deficit/Hyperactivity Disorder (ADHD) in a precise and specific manner.

**Methods:** The proposed pipeline will consist of: (1) a gamified web applications to curate videos of social interactions adaptively based on needs of the diagnostic system, (2) behavioral feature extraction techniques consisting of automated ML methods and novel crowdsourcing algorithms, and (3) development of ML models which classify several conditions simultaneously and which adaptively request additional information based on uncertainties about the data.

**Conclusions:** The prospective for high reward stems from the possibility of creating the first AI-powered tool which can identify complex social behaviors well enough to distinguish conditions with nuanced differentiators such as ASD and ADHD.

## Introduction

Roughly 17% percent of minors in the United States aged 3 through 17 years have a diagnosis of one or more developmental or psychiatric conditions [1], with the true prevalence likely being higher due to underdiagnosis in rural areas and for minority populations [2]. Unfortunately, timely diagnostic services are inaccessible to a large portion of the United States and global population due to cost, distance, and clinician availability. Digital phenotyping tools have the potential to shorten the time-to-diagnosis and to bring diagnostic services to more people by enabling accessible evaluations. While automated machine learning (ML) approaches for detection of pediatric psychiatry conditions have garnered increased research attention in recent years, existing approaches use a limited set of social features for the prediction task and focus on a single binary prediction.

Many psychiatric conditions affecting adolescents contain overlapping etiologies and phenotypic characteristics. A major difficulty preventing expansion of computational methods into simultaneous prediction of multiple related conditions stems from heavy similarities between their phenotypes, creating barriers to achieving specificity and precision. While some of the key overlapping and distinct features of these conditions are related to behaviors which can be automatically detected with ML methods, such as eye gaze patterns and facial emotion evocation, the majority are too complex for current ML techniques to classify precisely. For example, the degree to which a child enjoys participating in social games and interactions is one of the most salient behavioral features for ASD diagnosis [3], but building a classifier for such behaviors without overfitting is infeasible using current deep learning techniques and available datasets. By contrast, humans can naturally identify complex and nuanced behaviors by observing a subject. Crowdsourcing, or the use of distributed human workers towards a common goal, has the potential to bridge this gap by enabling rapid feature tagging of complex behaviors on-demand. While crowdsourcing has traditionally been used for public health studies and labeling ML training data, I plan to explore the incorporation of human labelers into the feature extraction process. The intuition behind the proposed paradigm is that while non-professionals may be unable to directly identify psychiatric diagnoses from videos, many can tag behaviors which are relevant to a diagnosis.

I propose to develop a novel paradigm for accessible and scalable multi-condition digital diagnostics of neuropsychiatric conditions by fusing traditional ML with novel human-in-the-loop crowdsourcing approaches. While this approach (Figure 1) can be applied towards classification between any set of psychiatric conditions, I will focus on Attention-Deficit/Hyperactivity Disorder (ADHD) and Autism Spectrum Disorder (ASD) to maintain feasibility. The approach will comprise of (1) a gamified web applications to curate videos of social interactions adaptively based on needs of the diagnostic system, (2) innovative behavioral feature extraction techniques consisting of automated ML methods and novel crowdsourcing algorithms, and (3) ML models which classify several conditions simultaneously and which adaptively request additional information based on uncertainties about the data. I will collaborate with Dr. Dennis Wall, who will provide domain expertise for pediatric developmental delays and methodological guidance for innovative biomedical data science solutions.

**Figure 1.**
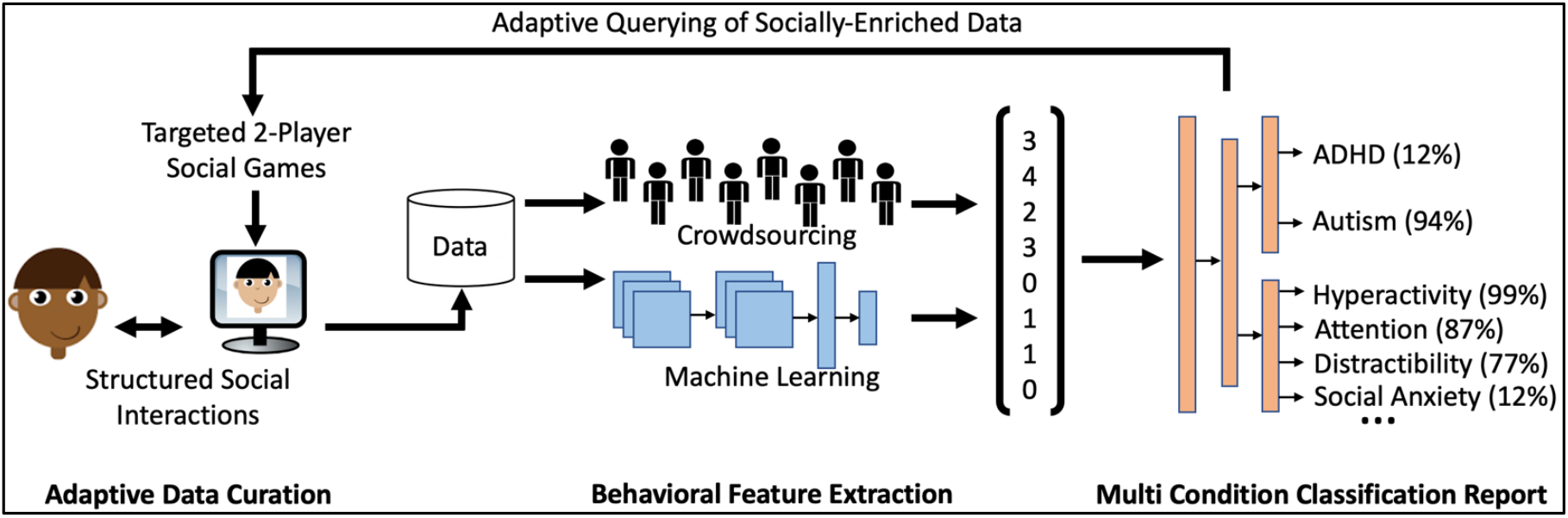
Overview of the proposed crowd-powered diagnostic system comprising adaptive gamified data curation, behavioral feature extraction by both crowd workers and computational workflows, and ML models for multi-condition classification which also output individual symptom estimates and dynamically query participants based on crowd ratings. Each of these three major steps are independent yet can be combined to produce a synergistic improvement in remote and accessible diagnostics for pediatric psychiatry.

I hypothesize that diagnostic ML models which incorporate both human-annotated features acquired through crowdsourcing (to generate a complex feature space with respect to social human behavior) and automatically extracted features (to provide objectivity when possible) will outperform models which use only automatically extracted features or only human-provided features, as there will likely be nonlinear interactions between features. This complex feature space will allow the classification model to simultaneously distinguish four possible outcomes: only ASD, only ADHD, ASD and ADHD, or neither condition. To support efficient and reliable feature tagging by workers, I will develop novel crowdsourcing algorithms for quantifying the behavioral tagging strengths and weakness of each worker. The algorithms will dynamically assign workers to tasks based on their tagging history. I will alter each video to provide privacy protections for the human subject while still allowing reliable tagging. To facilitate the acquisition of sufficiently structured data, I will develop a broadly accessible gamified web platform for curating socially enriched video and audio clips in a targeted manner. I will use active learning algorithms to adaptively query for additional data in cases where presence of a particular symptom is unclear from the current set of ML features and crowdsourced ratings. Each of these innovations (crowdsourcing algorithms, privacy-preserving video alterations, gamified social data capture system, and active learning algorithms to dynamically query needed data), while useful for the field of precision psychiatry individually, will be combined to create a novel diagnostic system with greater discriminative power than previously possible.

Psychiatric conditions are widespread globally across demographic groups and geographic boundaries. The prevalence of ADHD is 2.5% in children and 5% in adults [3]. The prevalence of ASD is roughly 1% [3]. About 50% to 70% of individuals diagnosed with ASD also have comorbid ADHD. Access to diagnostics, and therefore care, is limited for populations with low income or large geographic distances from clinicians. While diagnostic modalities based in biomarkers are promising, they can be inaccessible to underserved populations. By contrast, a large and rapidly expanding portion of the global population has access to digital devices. Since psychiatric conditions are fundamentally diagnosed from behaviors, digital methods to measure behavior have the potential to bring diagnostic services to populations that have been traditionally neglected in healthcare.

A psychiatrist’s diagnostic evaluation process involves identifying one or more conditions from a large set of possibilities defined by the DSM. However, current approaches to digital diagnostics tend to focus on binary predictions. A major bottleneck complicating the pursuit of multi-class psychiatric diagnostics is that behavioral conditions often have overlapping presentations (Table 1), severely complicating the use of purely automated methods. Additionally, each condition is heterogeneous, and all defining behavioral symptoms do not have to be present to warrant a diagnosis. Psychiatric conditions can either be comorbid (e.g., ADHD and ASD) or not (e.g., only ADHD or only ASD), creating a diagnosis space which scales combinatorically with each additional condition considered. For feasibility, I will only study 2 conditions to maintain a reasonably sized output space of 4.

**Table 1.**
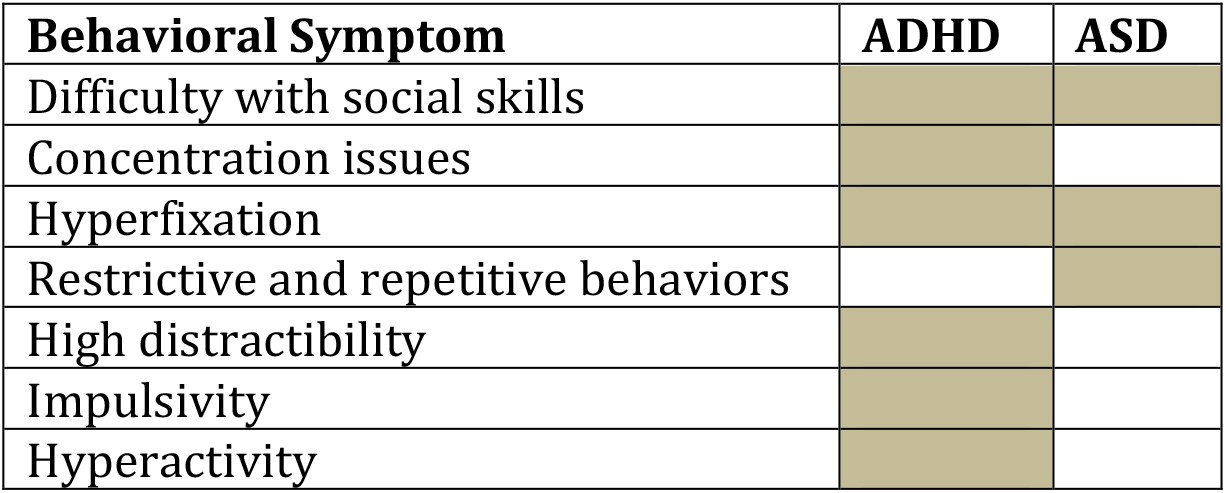
Overlap of a small subset of the core behavioral symptoms of ASD and ADH. Overlap is determined according to the DSM-V diagnostic criteria [3].

The proposed project involves the integration of multiple data modalities for its diagnostic tasks, including from ML and from crowd workers. In my prior work, I have worked with several sources of information such as facial emotion [4-5], body movements [6-7], audio streams [8], and crowd worker ratings [9-10], all used towards the singular goal of digital ASD diagnostics. For this proposal, I hypothesize that the complex and heterogeneous nature of the conditions which I plan to study requires multimodal data analysis to achieve a clinically acceptable level of performance, and this proposal will involve testing this theory.

### Related Work

The field of digital phenotyping is vast and broad. A non-exhaustive list of NIH-funded projects for developmental diagnostics include Guillermo Sapiro’s work (R01MH120093) developing active closed-loop data collection for gaze and motor features for ASD as well as ADHD [11-18], James Rehg’s work (R01MH114999) modeling nonverbal communication in atypical and typical development [19-20], Robert Schultz’s work (R01MH118327) involving diagnostic computer vision analyses of motor movements displayed in videos of dyadic social interactions involving children with ASD [21], and Dennis Wall’s work (R01LM013364) exploring the use of mobile games to acquire computer vision data for deep learning prediction of individual ASD-related behaviors [22-37].

In contrast to these inspirational funded efforts and others like them, I propose a approach to digital phenotyping that expands the possible feature vectors used to classify psychiatric conditions with complex and nuanced social features that only humans can identify using a novel “crowd-powered precision diagnostics” approach. I will collaborate closely with Dr. Dennis Wall on this work. The primary high-risk and high-reward differentiators from prior work are (1) the incorporation of a novel crowdsourcing pipeline into a precision diagnostic system to enable quantification of more complex social features, (2) the adaptive querying of the subject in question within a two-player game-based system using active learning algorithms which exploit crowdsourced responses, and (3) the differential diagnosis of ASD and ADHD simultaneously. Differentiators (2) and (3) would not be possible without (1). The addition of targeted crowdsourcing into the diagnostic process creates several technical challenges which I will address, including automating the preservation of privacy of human subjects, efficiently and intelligently quantifying the behavioral feature tagging ability of crowd workers, and creating algorithms for dynamically assigning workers to new data streams and tasks. While prior projects have attained successful performances >90% using purely automated deep learning approaches to differentiate ASD from neurotypical subjects [38], my preliminary data show that human-in-the-loop crowdsourced feature tagging of targeted behavioral features results in classification sensitivity, specificity, and accuracy >95%, even when privacy-preserving alternations are made to the video streams [39-41]. I hypothesize that incorporating both human observations, which are beyond the current and foreseeable abilities of ML, into the feature extraction process will provide enough social information for automated models to classify each condition using the same video data.

## Methods

### Overview

Achieving the precision required to distinguish between ASD, ADHD, both ASD and ADHD, or neither from videos of social interaction using ML at clinically acceptable levels requires a complex social feature space that is not necessarily impossible but highly infeasible with purely automated methods. In contrast, untrained human annotators can identify nuanced social features but are prone to error due to the subjective nature of the task. Combining features extracted by both non-expert human raters and computational programs can enable precise diagnostics and quantification of behaviors by creating a rich diagnostic feature space. There are several challenges to accomplishing targeted crowdsourcing in a precision health context, which I will address, including privacy preservation, quantifying crowd worker capabilities, and developing algorithms for matchmaking crowd workers with incoming data streams. The rich social feature space provided by crowdsourcing enables improvements to the other aspects of the digital behavioral diagnostics pipeline, including adaptive assignment of subjects to data collection games using active learning crowdsourcing metrics. While I will focus on ASD and ADHD in particular, the crowd-powered methods I will develop have the potential to benefit diagnostics for any condition primarily evaluated through behavioral observation.

## Gamified Data Curation

### Description

I will develop novel gamified social experiences to curate video data containing diagnostically rich information. These games will each impose the structure required to extract salient behavioral features which are comparable across subjects. Each game will involve 2 participants interacting virtually through both the game itself and socially through live video and audio. During gameplay, each participant’s camera and microphone will be turned on, and their video and audio will be displayed in a Zoom-style feed to the other participant. The video and audio feeds will be recorded during each session in addition to keyboard strokes and mouse movements.

Each game will correspond to a subset of targeted behaviors for data capture. Existing literature on “serious games” have documented the usefulness of certain games to capture behaviors related to psychiatric diagnostics, although these games are usually single player. One example is a Go/No-Go game, where the player presses the spacebar in response to a timed “go” prompt with the presence of auditory and visual distractors. This game has been shown to be a reliable estimate of attention, impulsivity, hyperactivity, and executive functioning when recording gaze behavior, response time, and correct reaction rate [43]. I will modify the game so that the “go” prompts are initiated by the social game partner rather than an automated computer, allowing for capture of socially relevant features. The field of “serious games” for assessment of psychiatric behaviors is vast, and I will therefore base all games on previously published literature. However, many behavioral features I will study will not be tied to a particular game but will rather be observable as a byproduct of the social interactions between participants (e.g., social anxiety).

One of 7 possible games will be administered each day. A complete list of games and corresponding behaviors that each game is designed to measure is shown in Table 2. Free-form conversation will naturally occur across all games. The design of the games will be conducted in consultation with a team of practicing clinical psychiatrists at the University of Hawai‘i School of Medicine Dr. Anthony Guerrero who is the chair of the Department of Psychiatry and who specializes in digital technologies for pediatric and adolescent psychiatry as well as Dr. Gerald Busch who is an Assistant Professor in the Department of Psychiatry who has experience with digital health solutions for psychiatry.

**Table 2.** List of previously validated data capture games which have successfully generated data relevant for distinguishing the targeted psychiatric conditions from neurotypical controls. Because the games themselves are not central to the innovation of this proposal, details of the gameplay can be found in corresponding references [5, 43-44]. *Free-form conversation will naturally occur across all games.

| Game Name | Targeted Behaviors |
| --- | --- |
| Go/No-Go [43] | concentration, impulsivity, hyperactivity, executive functioning, reaction time |
| AULA Nesplora [43] | process speed, motor activity |
| Plan-It Commander [43] | planning, organization |
| Braingame Brian [43] | working memory, cognition flexibility, impulsivity |
| Charades [5] | emotion evocation and recognition, restrictive and repetitive behaviors |
| Balloon Popping [44] | visual motor coordination |
| Spot The Eyes and Face [44] | eye contact, face gaze |
| Free-form conversation* | social anxiety, difficulty with social skills, speech delays, language narrative |

A minimum of 15 minutes of gameplay will be required each day, although participants may elect to participate for longer. To facilitate consistent data capture across possible computer, microphone, and camera configurations, a pertinent step for enabling comparisons across participants, a calibration program will be developed which will require each participant to align the camera’s zoom and their body position prior to each session. I will extensively test the calibration procedure prior to the study.

### Participant Recruitment and Management

I will recruit 100 participants formally diagnosed with ADHD, 100 with ASD, 100 with both ASD and ADHD, and 100 evaluated and confirmed to not have a socially related psychiatric condition, for a total of 400 study participants. I will recruit a population balanced by race, ethnicity, and gender. While formal and well-established methods to perform power calculations for ML analyses have yet to be established, most digital diagnostics studies for conditions such as ASD include less than 100 participants per class in binary classification [38]. I aim to maintain a similar sample size per diagnostic category. The digital social experiences will be delivered to study participants for 15 minutes each day for a 3-week duration, with a single game out of the 7 possible delivered each day. At least 3 distinct 15-minute sessions will be collected per game for each participant, allowing for comparisons across days for analysis of within-subject consistency.

Given the remote delivery of the data collection, a critical challenge will be to ensure that all participants will have a social partner when logging into the study system. I will ask participants to log in at a particular time each day, to be scheduled in advance of the first day of the study. I will host 10 separate time slots and 3 make-up timeslots every day of the study, and participants will be automatically matched with a partner during login.

### Evaluation

I will evaluate the data curation system for (1) compliance of participants with respect to the study procedures during each session and (2) global participation rates. To measure compliance, I will run computer vision face detection algorithms in conjunction with skeletal pose estimation using MediaPipe to ensure that each participant’s face, upper torso, and shoulders are fully visible. I will calculate the percentage of valid frames across sessions per participant. To measure participation across sessions, I will record the total number of sessions with both participants and the mean session time. In addition to these quantitative analyses, I will run qualitative pilot user studies to understand participant’s experiences about the data collection game process, including questions about the entertainment value provided by the games, the usability of the participant matching and scheduling system, and open-ended feedback.

### Feasibility

The technical aspects of the project are highly feasible, with modest development requirements compared to modern real-time computer gaming systems. The web server will be developed using the Django Python framework and hosted on an Elastic Cloud Compute (EC2) instance on Amazon Web Services (AWS), with extensive existing functionality and documentation existing for all technologies used. Extensive online code exists for implementing the video and audio chat features. A full-time developer, engineering/computer science student, or postdoctoral researcher can implement the entire system within the span of 4 person-months.

The primary challenge will be the recruitment and retention of 400 study participants, including the formal clinical validation of diagnosis for each participant. To help manage this recruitment effort, I will hire a full-time clinical research coordinator to recruit and manage participants. I will work with my clinical collaborators in the Department of Psychiatry to recruit in Hawai’i’s psychiatric clinics where my collaborators and their colleagues practice. I will supplement this recruitment with online recruitment using targeted advertisements on social media. I have discussed this recruitment plan and desired study size with my collaborators in the Department of Psychiatry, and we hold recurring monthly meetings to strategize about participant recruitment using both our existing access to several clinics in Hawai’i and online targeted recruitment. Additionally, my former mentor and collaborator Dr. Dennis Wall at Stanford University has access to hundreds of families with adolescent children diagnosed with ASD as well as comorbid ADHD.

### Potential Pitfalls and Mitigation Strategies

The proposed data curation game platform may lack qualities which would garner repeated participant engagement over a 3-week period, such as poor user interface design, poor design of the automated notification system, or poor entertainment quality of the individual games. To mitigate this risk, I will leverage my formal graduate training in human-centered design) to run several iterative design sessions. I will consult with my long-time mentor Dr. Terry Winograd from Stanford University, who is one of the founders of the field of Human-Computer Interaction, regarding proper implementation of the design process to maximize both user engagement and high-fidelity data collection. I will run several pilot studies to obtain both qualitative and quantitative measures of engagement prior to running the primary data collection study.

Other potential pitfalls are compliance and tardiness. I will run automated computer vision checks in real time to ensure participant compliance with camera calibration requirements. Another script will send automated text message and email reminders to late participants, assigning them to make-up sessions. If these mitigation steps fail or if recruitment is unsuccessful and there are fewer than 100 participants with valid data per diagnostic category, the study can still be successful with as low as 20 participants per class, as ML studies with around 20 participants per diagnostic category have frequently been published in the field [38].

### Novel crowd-powered and traditional ML-based feature extraction *Description*

#### Description

I will create two pipelines for converting raw video and audio data into interpretable feature vectors which quantify social behavior relevant to ADHD and ASD diagnostics. For behaviors which can feasibly be quantified using computational methods, I will use existing toolkits. For highly complex and nuanced social behaviors which are beyond the scope of current ML tools, but which are highly relevant towards psychiatric classification, I will use a novel crowdsourcing pipeline to match crowd workers to labeling tasks.

I will run automatic feature extraction for behaviors potentially related to diagnoses such as percentage of total conversation time contributed by the participant, eye gaze patterns during the gameplay including proportion of gaze directed towards the game versus the live video feed of the other participant, vocal prosody and intonation during conversation, natural language processing analysis of the content of the conversation after converting raw audio to text using speech-to-text programs, and breaks in task flow as measured by pauses in game-related keystrokes and mouse movements. Extracted information will be stored for each frame at a sampling rate of 5 frames per second. As depicted in Figure 2, each of these features will be concatenated into a temporal feature vector and used to train a time series deep learning model such as a long short-term memory (LSTM) recurrent neural network or an attention-based model (e.g., Transformers). Existing Python libraries enable the proposed automatic behavioral feature extraction. For eye gaze, I will use the OpenFace and MediaPipe Python libraries. For facial emotion recognition, I will use Amazon Rekognition, an AWS service which provides recognition of disgust, happy, surprise, anger, confusion, calmness, and sadness in addition to other relevant facial features such as whether the eyes and mouth are open. In the audio domain, pitch will be extracted using the CREPE library, and waveforms will be processed using the librosa library.

**Figure 2.**
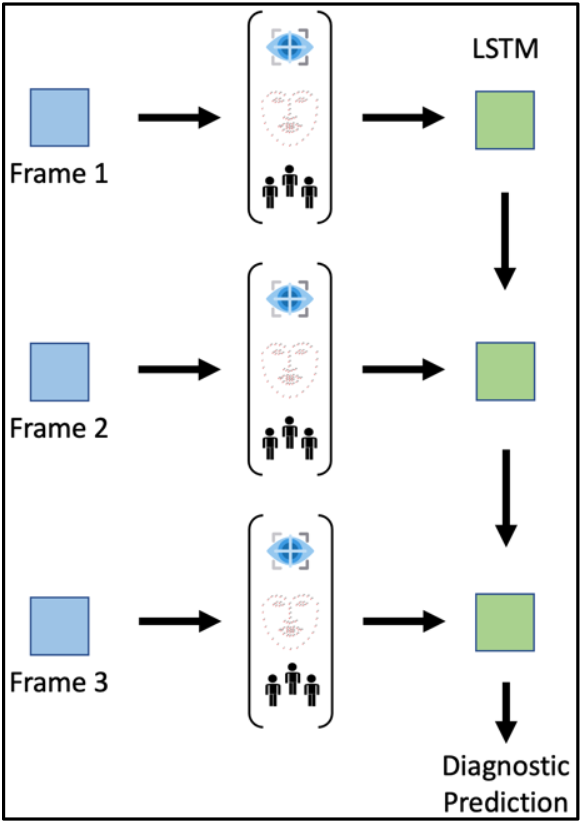
Feature extraction and quantification of behaviors relevant to neuropsychiatric diagnostics.

**Figure 3.**
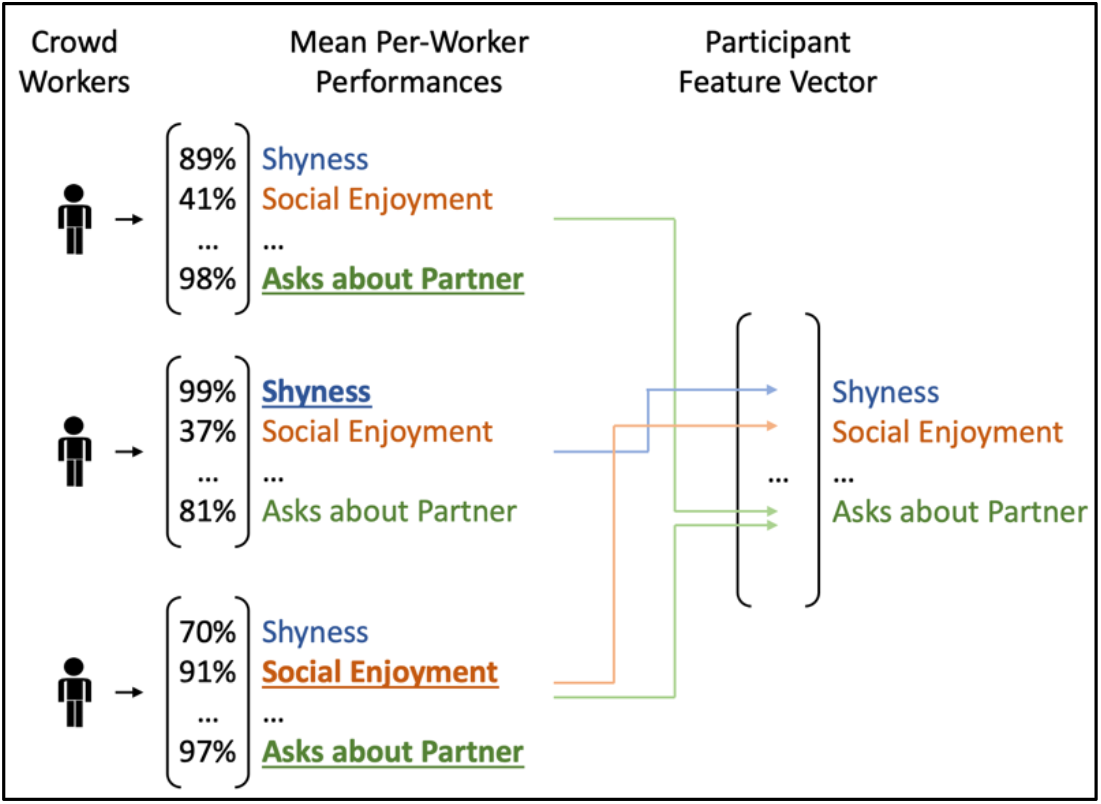
Crowd worker assignment to labeling tasks. Each crowd worker will only be asked to label those features for which they agreed with clinicians during a worker profiling step performed prior to the primary study.

For complex social features beyond the scope of automated ML-powered computational processing, I will deploy a novel crowdsourcing framework consisting of a crowd worker profiling phase followed by a study data tagging step. In the first phase, I will post 20 tasks on Amazon Mechanical Turk, each presenting a video acquired through pilot testing of the gamified social data collection platform followed by a series of multiple-choice questions corresponding to items from the diagnostic criteria for ADHD and ASD as defined by the DSM-V. Each task will correspond to a separate video, and there will be 4 videos per diagnostic category used to quantify worker abilities. Worker responses will be compared against gold standard ratings provided by my collaborators in the Department of Psychiatry at the University of Hawai’i. Crowd workers who align with the ratings of clinical experts on at least 1 behavioral feature, where alignment is defined as less than 1 categorical ordinal deviation per 2 videos, will be recruited to label the final study data from 400 participants. Recruited workers will only label those features for which their alignment with clinicians was demonstrated during the profiling phase.

To ensure that the annotation task is manageable for crowd workers, each 15-minute video will be segmented into five 3-minute clips. During the profiling phase, crowd workers will be compensated $0.50 per 3-minute video segment rated.

Workers who are selected to continue rating videos in the primary portion of the study will be compensated $0.05 per question answered per video segment, with the opportunity of a bonus of $0.05 per question if the answer aligns with the clinician ratings for that question. These payment rates are consistent with practices in crowdsourcing research studies in the field of human-computer interaction, and my preliminary studies have shown that the retention rate for this level of compensation is above 90% [9, 40-41].

Participants whose videos will be shared for the 20 crowdsourcing tasks used to filter workers will be contacted by the study team to have a thorough discussion about the planned use of those videos. Workers who are qualified to rate the remaining videos for one or more questions will be required complete HIPAA training and CITI training and will be required to encrypt their laptops using whole-disk encryption. These workers will be added to the IRB protocol and will become official members of the study team after thorough training.

While I will require participants in this study to consent to sharing videos with crowd workers who have undergone thorough training, clinical translation of this diagnostic system will require a more scalable approach which is sensitive to privacy concerns. I will experiment with privacy-preserving alterations to the curated videos to obfuscate identifiable information from the videos while not degrading the feature tagging performance of workers. Examples include pitch shifting the audio, which will allow workers to understand the content of the speech, and pixelating the video, which will obscure the participant’s background and face but would still allow workers to observe body movement patterns. I will measure the amount that each privacy-preserving mechanism degrades answers to each question.

A valuable byproduct of this process will be the generation of large behavioral multimedia datasets for ML of complex social features, enabling improved AI modeling of human behavior more broadly. With explicit permission from study participants on a per-video basis, I will package and publish the collected data into novel computer vision, audio, and NLP datasets for ML. These labeled datasets will be released publicly, providing a steppingstone towards improved automated methods for quantifying complex human behavior.

### Evaluation

To evaluate the crowdsourcing pipeline, I will compare the performance of crowd workers prior to being recruited with the performance after recruitment. My preliminary data show that crowd workers who answer similarly to clinicians during filtering continue to perform in a similar manner on new, unseen videos. I will also measure crowdsourcing metrics such as latency to starting a task, inter-rater reliability, any decline in performance with increased ratings, and completion rate for all assigned tasks. To evaluate the privacy-preserving mechanisms, I will randomly assign each worker to a single privacy condition per video, only asking them to rate the unaltered videos after the ratings for the privacy condition have been provided. I will measure the mean deviation from clinician answers per privacy condition for each question.

### Feasibility and Preliminary Data

I have conducted a series of preliminary studies testing the use of crowdsourcing for precision behavioral health, demonstrating that while there is a high degree of variability in crowd workers’ innate ability to rate complex social behaviors in unstructured home videos [9], there exists a small fraction of crowd workers on platforms such as Amazon Mechanical Turk who consistently rate in alignment with licensed clinical experts [39-40]. In one study, I demonstrated that a group of 40 crowd workers filtered from an original pool of >1000 workers were able to rate behaviors which, when fed into a classifier trained on clinician records, achieved an AUROC of 0.9904 for one set of features and 0.9872 for another feature set [41]. My experience receiving approval from University IRBs and Data Privacy Offices as well as obtaining consent from families to share their videos with crowd workers mitigates any risks about this novel process.

After applying privacy-preserving modifications to the videos, such as pitch shifting the audio downwards and using face detection to box out the child’s face, the performance of the model remained above 0.95 for both AUROC and AUPRC [41]. These results, while promising for the binary task of autism prediction, are likely to degrade when expanding include ADHD. These studies provide strong evidence to support the proposed worker matching procedure which will enable the more nuanced feature space required for multi-condition classification. My prior experience developing automated pipelines for managing crowd workers will help streamline the development of the crowd management scripts.

The feasibility of the automatic feature extraction steps comes from the existing packaging of the required functionalities into Python libraries and the high documented performance of these tools. All ML-powered feature extractors I have used are well documented.

### Potential Pitfalls and Mitigation Strategies

While this never occurred during collection of my preliminary data, a potential pitfall is that some questions may have no workers who consistently rate in accordance with clinicians. If this occurs, then that question will be removed from any further components of the study (i.e., removed as a feature for the diagnostic classifier). Another possible pitfall is that recruited workers will not complete their assigned ratings. During my preliminary studies, I observed >90% retention when workers were compensated at the level I am proposing. To handle the workers that drop off anyways, I will recruit three times more crowd workers than minimally required for the study, which is easily feasible with a crowdsourcing platform with a large user base such as Amazon Mechanical Turk.

When verifying the automatically extracted computational features by hand, it is possible that some features will be incorrect. Computational feature extraction approaches are not perfect and are not necessarily robust to unforeseen conditions (e.g., dim lighting, obfuscation of certain body parts, and unfamiliar accents). If any feature is consistently unreliable across several participants, then I will remove that feature from the study. There are enough features that if some do not work, the study can proceed.

### Multi-condition diagnostics with adaptive input querying

#### Description

I will develop deep learning models for multi-label classification of ADHD and ASD, which can emit 4 possible outcomes: (1) ADHD, (2) ASD, (3) ADHD and ASD, and (4) neither condition. The models will also output the behavioral characteristics that led to the final classification decision by producing the percent confidence of each behavior as derived from both crowd workers and automated computational models. This will involve synthesizing multiple sources of inputs and communicating the result to the end user in a manner which is understandable by the patient or caregiver. The confidence scores will enable the model to adaptively request more data from the patient and to be specific about which types of data are needed.

To derive an interpretable quantification of each behavioral feature, I will collect clinical categorical ratings of each behavior by licensed psychiatrists at the University of Hawai’i at Mānoa. I will compensate the psychiatrists for their service. I will use the mean of the crowd worker responses as a baseline method for deriving the interpretable quantification of each behavior. While this method could be sufficient, it is possible that crowd workers have varying levels of rating abilities depending on qualities of the video and qualities of the crowd workers themselves. I will therefore explore the use of the crowdsourced ratings themselves combined with crowdsourcing metrics derived from worker performance and the computationally generated behavioral features as collective inputs into an ML model for each behavior. Such metrics will include time spent by each worker providing the annotations for the video, worker rating history for each question, and variability in the worker’s answers across videos and within a particular video. I have previously shown that these crowdsourcing metrics and similar metrics have predictive power in a crowd worker’s annotation quality [10]. I will test whether the ML model is a better predictor than the crowdsourced ratings alone. The loss function for the ML model for individual behaviors will optimize with respect to the mean clinician rating per behavior.

To model the multi-label classification problem, I will create separate binary classifiers for ASD and ADHD. Each model will be optimized separately. Using a sigmoid activation function for each independent classifier, the classification system will output a probability score for each diagnostic possibility as well as each of the behaviors defined by the DSM-V which will be quantified by the system.

Using the output scores of the deep learning model, I will develop an active learning system which queries for additional data from the user in a targeted manner by suggesting the next game for the participant to play. For each participant, the algorithm will measure the confidence score of each behavioral symptom and produce a list of games for the user to play sorted by the classifier’s mean uncertainty of the behavioral symptoms each game is designed to curate data for.

Classifier uncertainty will be measured by the entropy of each classifier’s output vector. Because neural networks are inherently uncalibrated, I will apply a method published by Kuleshov et al [42]. based on isotonic regression to calibrate the probability estimates prior to measuring uncertainty.

#### Evaluation

I will evaluate the diagnostic ML model using balanced classification metrics including Area Under the Receiver Operating Characteristic (AUROC) curve, Area Under the Precision-Recall Curve (AUPRC), balanced accuracy, precision, recall (sensitivity), F1-score, and specificity. Performance and confidence intervals will be derived through Monte Carlo cross validation, with each data split consisting of 300 participants in the training set, 50 participants in the validation set, and 50 participants in the test set. All splits will contain balance with respect to the 5 diagnostic classes, age, gender, race, and ethnicity.

To evaluate the effectiveness of the active learning querying system, I will run post hoc simulations comparing random selection of new data against targeted requests using active learning. I will train the classification system with 12 sessions of data and hold out the remaining 9. I will plot the performance of each metric against the number of additional samples acquired using both active learning and random selection of data segments.

#### Feasibility

The feasibility of deep learning models relies on the underlying data used to train them. Deep learning has the capacity to learn any discriminative function provided a large enough model and adequate computational power to train a large model. My institution has provided me with a dedicated Nvidia v100 GPU node and a dedicated Nvidia RTX5000 for computationally intensive research. In addition, the Hawai’i Data Science Institute has shared computing resources consisting of 346 nodes (8,500 cores) with a total of 63.19 TB of RAM, 120 GPUs and more than 1 PB of storage. These resources are free to use for University of Hawai’i labs. Collectively, these resources are more than sufficient to train deep learning models for the proposed dataset size.

I have previously trained deep learning models for making a binary prediction of ASD (Table 3). Although these models each used a single data modality (audio, facial emotion expression, or eye gaze), the performances were on par with prior literature [38]. I hypothesize that incorporating additional modalities will not only allow for increased performance within a single class but will enhance discriminative power across diagnostic categories.

#### Potential Pitfalls and Mitigation Strategies

It is possible that the large number of features used to train the deep learning classification models will be overfit to the training set, as a dataset of 400 samples (of which about 75% would be in the training set) is relatively small for ML and is unlikely to capture all of the intricacies of social behavior which can be expressed with the feature space. If this happens, I will run feature selection and dimensionality reduction algorithms to reduce the number of features used in the model to a minimum viable set and to summarize the feature space in a low-dimensional manner, respectively. The feature selection will enable interrogations into which features are most useful in the differentiation of each condition.

## Data Availability

There are no data associated with this Research Protocol.

